# The social experience of participation in a COVID-19 vaccine trial: Subjects’ motivations, others’ concerns, and insights for vaccine promotion

**DOI:** 10.1101/2020.12.30.20249051

**Authors:** Emily Wentzell, Ana-Monica Racila

**Affiliations:** Department of Internal Medicine, Carver College of Medicine, University of Iowa Hospitals and Clinics, 200 Hawkins Drive, Iowa City, IA 52242 USA

## Abstract

**Background:** Vaccine hesitancy could undermine the effectiveness of COVID-19 vaccination programs. Knowledge about people’s lived experiences regarding COVID-19 vaccination can enhance vaccine promotion and increase uptake.

**Aim:** To use COVID-19 vaccine trial participants’ experiences to identify key themes in the lived experience of vaccination early in the vaccine approval and distribution process.

**Methods:** We interviewed 31 participants in the Iowa City, Iowa US site of the Pfizer/BioNTech COVID-19 vaccine phase 3 clinical trial. While trial participation differs from clinical receipt of an approved vaccine in key ways, it offers the first view of people’s lived experiences of potentially receiving a COVID-19 vaccine. The trial context is also useful since decision-making about vaccination and medical research participation often involve similar hopes and concerns, and because the public appears to view even approved COVID-19 vaccines as experimental given their novelty. Semi-structured interviews addressed subjects’ experiences, including decision-making and telling others about their trial participation. We analyzed verbatim transcripts of these interviews thematically and identified common themes relevant for vaccination decision-making.

**Results:** Participants across demographic groups, including age, sex/gender, race/ethnicity, and political affiliation, described largely similar experiences. Key motivations for participation included ending the pandemic/restoring normalcy, protecting oneself and others, doing one’s duty, promoting/modeling vaccination, and expressing aspects of identity like being a helper, career-related motivations, and support of science/vaccines. Participants often felt uniquely qualified to help via trial participation due to personal attributes like health, sex/gender or race/ethnicity. They reported hearing concerns about side effects and the speed and politicization of vaccine development. Participants responded by normalizing and contextualizing side effects, de-politicizing vaccine development, and explaining how the rapid development process was nevertheless safe.

**Conclusion:** These findings regarding participants’ reported motivations for trial participation and interactions with concerned others can be incorporated into COVID-19 vaccine promotion messaging aimed at similar populations.

## 1. Introduction

Vaccines will play a key role in stemming the COVID-19 pandemic, particularly in sites like the US characterized by inadequate behavioral mitigation strategies and politicization of public health measures [1, 2]. However, vaccine hesitancy threatens to undermine the effectiveness of this approach [3]. A recent US survey found that 27% of respondents anticipated probably or definitely not getting a free, approved COVID-19 vaccine. Attributes including political affiliation, race/ethnicity, and occupation mediated reported hesitancy, with higher percentages of self-identified Republicans, Black adults, and essential and healthcare workers anticipating hesitancy or refusal [4].

Vaccine hesitancy is not a fixed trait but a calculation someone makes about the perceived risks of vaccination versus disease in a specific social and structural context [5, 6]. Hesitancy is strongly mediated by experiences of group membership and values [7, 8]. It can also be a response to prior harms and ongoing inequalities, like anti-Black medical racism and legacies of colonization [9, 10]. Accordingly, hesitancy is best countered through structural measures ensuring transparent safety monitoring, equitable distribution, and affordability; and social measures, including peer role modeling and media campaigns and interpersonal communication efforts which address individuals’ context-specific concerns [11-14].

Knowledge about the context-specific factors affecting COVID-19 vaccine decision-making is thus necessary for increasing vaccination acceptance [15, 16]. Over half of Americans who anticipated hesitance toward a future approved COVID-19 vaccine voiced concern about side effects, politicization of the development process, government failure to ensure safety and effectiveness, and vaccine novelty [4]. It is crucial to determine whether these remain key concerns in the context of actual rather than hypothetical vaccination.

Qualitative research with vaccine trial participants can generate actionable insights for enhancing vaccine acceptability [17]. Thus, COVID-19 vaccine trials offer a useful early site for generating such knowledge. Phase III trial participation differs from a typical immunization experience in that participants might receive placebo or an ineffective vaccine candidate and that fear of adverse effects might be greater for experimental vaccines. Nevertheless, trials are an invaluable comparative setting because similar factors motivate people’s decision-making regarding medical research participation and vaccination, including: efforts to be a good, caring person or citizen [10, 18]; to demonstrate belonging in or care for a social group [7, 19]; and to protect individual, family or community health [20-22]. Trial participants’ experiences are especially relevant for anticipating future responses to COVID-19 vaccines, since the public appears to view even approved COVID-19 vaccines as “experimental” given their novelty [4].

Here we report findings from interviews with participants in a US phase III clinical trial of the Pfizer/BioNTech COVID-19 vaccine. To discern themes useful for vaccine promotion, we aimed to identify subjects’ stated motivations for participating, the content of the positive and negative responses to their participation they reported hearing from others, and participants’ responses to others’ concerns.

## 2. Methods

We interviewed subjects in the Iowa City, IA, USA site of Pfizer/BioNTech COVID-19 vaccine trial about their social experiences of trial participation. Vaccine trial staff at the University of Iowa Hospitals and Clinics (UIHC) assisted in qualitative study recruitment. Trial staff gave all subjects an information sheet describing our study at their second immunization visit and asked if participants would like their names and contact information shared with the qualitative researchers. To aid in achieving a demographically similar sample to that of the vaccine trial, trial staff forwarded interested participants’ occupations, characterized as “healthcare” or “other.” Qualitative study staff contacted interested trial participants as we received their information, focusing on non-healthcare workers later in recruitment to ensure occupational balance.

Each participant engaged in one semi-structured phone interview with one of two researchers experienced in this methodology. Interviews were audio-recorded, lasted up to one hour, and averaged 33 minutes. Interview guides were designed according to best practices, including building rapport by beginning with low-stakes topics and soliciting concrete examples of lived experience [23]. Questions addressed three interrelated domains of experience: reasons for enrolling in the vaccine trial, social experiences of enrollment and participation, and understandings of vaccine and clinical trial science. Interviewers asked personalized follow-up questions to solicit full narratives regarding the experiences that participants described. Interviewees quoted below are identified with a participant number; readers can find the demographic information of each numbered participant in Table 1.

**Table 1.** Demographic Characteristics of Quoted Interview Participants

| *Participant ID* | *Gender/Sex* | *Age* | *Race/Ethnicity* | *Occupation* |
| --- | --- | --- | --- | --- |
| P1 | Female | 45-54 | White | Healthcare |
| P2 | Female | 18-24 | Hispanic/Latinx | Other |
| P3 | Female | 35-44 | White | Other |
| P4 | Male | 45-54 | Black | Other |
| P5 | Female | 45-54 | Hispanic/Latinx | Other |
| P6 | Male | 45-54 | White | Other |
| P7 | Female | 25-34 | White | Healthcare |
| P8 | Female | 45-54 | White | Other |
| P9 | Female | 45-54 | White | Other |
| P10 | Male | 45-54 | Hispanic/Latinx | Healthcare |
| P11 | Female | 35-44 | White | Other |
| P12 | Male | 18-24 | White | Other |
| P13 | Male | 45-54 | White | Other |
| P14 | Female | 45-54 | White | Other |
| P15 | Female | 45-54 | White | Other |
| P16 | Male | 55-64 | White; Hispanic/Latinx | Healthcare |
| P17 | Male | 55-64 | White | Other |
| P18 | Female | 18-24 | White | Healthcare |
| P19 | Male | 55-64 | White | Other |
| P20 | Male | 45-54 | East Asian | Other |
| P21 | Female | 35-44 | White | Healthcare |
| P22 | Male | 35-44 | White; Hispanic/Latinx;<br>South Asian | Other |
| P23 | Male | 55-64 | East Asian | Other |
| P24 | Female | 55-64 | White | Healthcare |
| P25 | Male | 18-24 | Hispanic/Latinx | Other |
| P26 | Female | 25-34 | White | Other |
| P27 | Female | 55-64 | White | Healthcare |

We analyzed verbatim interview transcripts thematically [24]. We incorporated both deductive and inductive approaches to identify themes present in the relevant literature as well as novel themes [25]. Author 1 coded the transcripts for these themes using the qualitative research software NVivo [26]. Author 2 double-coded 25% of the interviews (n=7) to check for agreement and reliability. We held consensus meetings throughout this process to identify themes and develop and refine codes. We concluded the interviewing process after achieving code saturation and meaning saturation for key themes [27].

### 1.1 Funding and Ethics

This research was funded by a University of Iowa Arts and Humanities Initiative grant.

The University of Iowa IRB deemed this project exempt (UI IRB ID# 202008019). All vaccine trial participants received an IRB-approved exempt information sheet describing the study aims and methods, and noting that the studies were separate and that people’s decisions regarding interview participation would not affect their vaccine trial participation. Interviewees were not compensated for participating in this qualitative study, although all vaccine trial participants were compensated. Qualitative study subjects gave verbal consent to be interviewed and audio-recorded.

## 3. Results

We interviewed 31 vaccine trial participants in September-November 2020; the final 5 interviews occurred after news of the Pfizer vaccine’s efficacy but before its Emergency Use Authorization from the US Food and Drug Administration. Iowa did not have a state mask mandate during this time and community spread of COVID-19 was high and uncontrolled [28].

Qualitative study participants’ demographics (Table 2) reflected those of the vaccine trial subjects. Themes and experiences participants voiced were remarkably consistent across demographic categories.

**Table 2.** Self-reported demographic characteristics of participants.

|  | Total (N = 31) |
| --- | --- |
| *Gender/Sex* |  |
| Female | 17 |
| Male | 14 |
| *Age* |  |
| 18-24 | 5 |
| 25-34 | 3 |
| 35-44 | 5 |
| 45-54 | 12 |
| 55-64 | 6 |
| *Race/Ethnicity\** |  |
| White | 23 |
| Hispanic/Latinx | 7 |
| East Asian | 2 |
| Black | 1 |
| South Asian | 1 |
| *Healthcare Worker* |  |
| Yes | 12 |
| No | 19 |
| *Native Iowan* |  |
| Yes | 18 |
| No | 13 |

\*Some participants identified with more than one racial/ethnic category, so n>31.

### 3.1 Motivations for trial participation

Participants overwhelmingly said they wanted to support vaccine development in order to help end the pandemic and facilitate what many described as a return to “normalcy.” In a typical comment, P1 noted, “If the vaccine works, we’re probably back to normal.” Similarly, P2 hoped her participation “gets us one step closer to going back to normal. Once we’re able to figure out this vaccine and once we’re able to mass produce it and get it out to people, I’m just hoping that this [pandemic] will go away.”

Participants also frequently voiced a longstanding desire to help others, which trial participation facilitated. Many identified as helpers, like P3 who explained, “I’m one of those people, I want to help people if I can.” Others characterized the desire to help as innate, as exemplified by P4’s comment that, “it should just be in mankind’s nature to want to help.” Participants often described the trial as an opportunity to do their “civic duty” or be a “good citizen.” Some saw doing this duty as everyone’s job, like P5 who noted, “I think everybody should contribute to a positive outcome, and I thought this was my way of contributing.” Others attributed their desire to help others to their life experiences or histories. For instance, P6 explained that he came from a long line of veterans and like his Normandy-survivor uncle used to say, “Somebody has to be the first one out of the plane.”

The ability to help others on a societal level featured in many participants’ justifications of vaccine trial enrollment to concerned others. For example, P6 quoted above recounted explaining his trial enrollment to a vaccine hesitant co-worker as follows: “I said, ‘I’m doing this for your kids and your grandkids. That’s what this is about. This isn’t about something to benefit me, it’s to benefit those who come next.’ Once we talked about it that way, then she understood that.”

Many participants felt called to help society via trial enrollment because they saw themselves as qualified to do so. They often described themselves as “healthy” enough to withstand participation, like P7 who noted, “I deal with anxiety and depression, but I’m otherwise super healthy. So I wasn’t concerned about getting sick or having a bad reaction from the vaccine.” Others thought their life circumstances made them suited to participation. This included some frontline workers, like teacher P8 who said, “it seemed like you’d want somebody that’s out there in person with the kids, that that’s somebody that you would want in the study.” P9 believed her lack of caregiving responsibilities made her especially eligible. She remembered being “impressed” with the “bravery” of a mom participating in an earlier trial and thinking, “Well, now I’m not married. I don’t have any kids. So if something bad goes wrong, I felt it was of a manageable risk, and I just wanted to do what I could to help.”

Participants mentioned the ability to represent their demographic group in the trial as an important motivator. This desire was commonly voiced by participants who identified as members of racial or ethnic minority groups that had been disproportionately affected by COVID-19 in the US. For instance, P10 said he felt the “need to make sure we had minorities represented in some of these clinical trials. And being also Hispanic, and having had a firsthand experience with the outbreak in the meat packing plant, where a lot of the Hispanic population ended up here at the University of Iowa. That also played an important role in some of my decision making.” P4 similarly stated, “me being a Black man, I wanted to be a part of something to try to help figure out a disease, a virus, that has really taken hold of my people.”

Women participants across racial/ethnic categories sometimes said they felt the need to represent their sex in the vaccine trial. P9 noted that, “medications and drugs are not tested on women enough. We metabolize things differently, and so I wanted to participate.” P11 thought that her sex, age, and reproductive choices together enabled her to provide needed demographic representation, since “…being somebody of my age and biological sex, there aren’t a lot of women who are not currently trying to get pregnant or get pregnant in the future who qualify.” Men as well as women of all ages noted their ability to represent their age cohorts. For instance, P12 said, “They probably needed young participants, so it’s worth putting our bodies on the line more than waiting for other people to.”

Participants overwhelmingly viewed their participation as support for the enterprise of science. Interviewees frequently identified “supporting” or “believing in” science and vaccines as key to their self-identity and participation. In a representative comment, P13 stated, “I just believe in science.” Several others were excited to participate in what they considered an unprecedented scientific achievement. As P14 explained, “I wanted to get in the study because of science, because this is groundbreaking science.”

Many participants worked in medical or scientific fields and described that as part of their ethos rather than just a job. P15 explained that, “working in public health, I think I have to be a champion for causes like this. Practice what I preach.” Similarly, P16 noted, “I do biomedical research, and whenever there is an opportunity to participate, I try to participate. I feel it’s my responsibility as somebody who enrolls people to do clinical research, to do the same, when somebody else is trying to advance science.”

Desire to support science led participants to engage in a range of activities beyond the COVID-19 vaccine trial. Several had donated blood, enrolled in cadaver donation programs, or participated in other health research. This included P17, who stated he was born with a cleft palate and had contributed to “great strides” in treatment though lifelong research participation. He explained, “I was kind of a Guinea pig from the get-go on that in terms of surgical procedures and that kind of thing… I’ve had probably close to twenty surgeries and they now have it down to five or six for the same results.” Another interviewee’s narrative reflected the combination of a general desire to help with the goal of supporting medicine. P6 expressed his gratefulness to UIHC staff for saving his wife’s life after an aneurysm, and reported wanting to help medicine advance even “if all I did was pick up cigarette butts outside the university.”

That same trust in science and its institutions made interviewees feel safe participating in the COVID-19 vaccine trial. For example, P18 explained, “I don’t assume to know everything about vaccines or vaccine research in general, but I feel like even though it has been accelerated, the process, I still feel like there is a lot of guidelines to make sure that things are safe.” Participants also relied on their own knowledge of the process to frame trial participation as safe. Several mentioned the phase III status of the trial to support claims that they themselves faced little risk. For instance, P19 noted, “I guess it was already in the phase three, so hats off to people that were in phase one and two, because they took the risk.”

Participants across the political spectrum frequently described supporting science through vaccine trial participation as a means to counter the harms of politicization of the nation’s COVID-19 response as well as public skepticism about vaccines and scientific integrity. P12’s “disgust of the fact that it’s politicized” encapsulated the common sentiment that, as P6 explained, “I believe there’s a time and place for politics and medicine isn’t one of them.” In a typical comment, P20 noted, “I think that people need to trust science and learn from authority, from scientists. And I don’t think politics and the other political movement should be involved in vaccine acceptance or usage or trial or decisions.”

In response to this perception that pandemic politicization fueled vaccine hesitancy, many participants also hoped to serve as a role models, inspiring others to receive an eventually-approved vaccine. P3 summed this goal up by saying “I feel like I should be a PSA to counteract the screaming headlines you see every now and then. I feel like it’s partly my responsibility to tell people, ‘Hey, I’m a participant. I’m not sure but I think I got the real thing. This is all that happened to me, almost nothing. Please, tell your friends, tell your neighbors, don’t be scared of the vaccine when it comes out.” Several participants with similar goals shared their experiences on social media. P21 explained, “I posted it on Facebook. Because my husband and I are both in the trial and hey, we are doing this. I felt like it’s important that people see” to inspire them into “getting the vaccine when it comes out.”

Many specifically hoped to counteract “anti-vax” messaging. For instance, P13 noted the presence of “a lot of anti-vaxxers” at both ends of the political spectrum, and said, “I don’t believe in it. And I think it’s silliness and I think it’s irresponsible to all of us. So that’s why I’m pushing that agenda a little bit, letting people know that I’ve got [the trial vaccine] and I’m safe.” Similarly, P22 said that sharing his experience was meaningful because, “I’m also pretty committed to doing what I can, when I can to help combat anti-vaxxer sorts of mentality.”

Some participants hoped their participation could demonstrate vaccine safety specifically for members of the groups with which they identified, especially racial or ethnic minority groups. For example, P22 identified as South Asian and said he hoped to lead by example for this and other minority “ethnic groups,” “some of whom have been deliberately mistreated with respect to vaccination in the past.” He hoped his example would make people “a little more comfortable with getting vaccinated, and trusting science and good medical advice and NIH and CDC and all of that.” Others hoped to allay concerns within their age cohort, like P13 who wanted “to show that people in my age group don’t have to be worried about it, that I have a good experience with it. And so then they should feel like they would have a good experience with it.”

In contrast, very few interviewees characterized their participation as “private” information to keep to themselves or within their immediate families, and some others shared it but less widely than the publicizers discussed above. Those who selectively shared information tended to tell only people they expected would be supportive. They avoided disclosing participation to people - often politically conservative relatives - who they thought might be opposed to COVID-19 mitigation measures. For example, P18 said, “I told my family and my friends and some people in med school with me because, I don’t feel like anyone in that group is really going to be dissenting with my choice I haven’t told anyone in my extended family where people tend to be more conservative when it comes to COVID and everything, a little bit more distrusting. So I pretty much only told people who I assumed would have similar opinions to what I have.”

In addition to seeking the societal-level public health benefits discussed above, some participants were highly motivated by the hope of receiving a potentially effective vaccine. Some focused on the desire to protect themselves from COVID-19. P23 noted, “I usually don’t participate any other trials, but this one I decided [to] because I really want to be vaccinated.” Many who voiced that desire framed trial participation as a win-win situation in which they might receive a potentially effective vaccine instead of placebo, and in any case would be contributing to a solution. For instance, P21 explained, “working in healthcare I definitely plan on getting the vaccine when it comes out. And I figured somebody has to join the trials in order to get to the point where it can be released even in a higher risk situation, but the sooner I can get access potentially to the vaccine the better.”

Participants frequently said they hoped to receive a potentially effective vaccine in order to protect vulnerable people in their lives. For example, P4 stated, “I felt like if it kept me from getting it, I wouldn’t be able to give it to my wife or my children.” P24 noted, “I knew I would have a chance of getting a real vaccine before everybody else and as a caregiver for an elderly person, that would be a good thing.”

Very few participants identified compensation as a reason for participating in the vaccine trial. Five of the six who did were in their 20s. One such participant, P2, described herself as “broke” and gave her reason for participation as “the money.” She also described research participation in transactional terms, noting, “I don’t really have any side effects, but if they come, it’s just part of the research. That’s why they’re paying me.” Conversely, with one exception, older participants who mentioned compensation either downplayed its importance or expressed ethical concern. Many volunteered that they were not motivated by money. For example, P9 recounted that when she called the trial to volunteer “they were like, ‘You will be compensated,’ and I was like, ‘Well that’s not why I’m doing it.’”

### 3.2 Others’ concerns and participants’ responses

Most participants reported receiving supportive responses from people they told about their trial participation, such as being thanked. However, most also reported hearing concerns. The most common concern involved negative side effects. In a typical statement, P15 said, “So, they just ask about my experiences and, ‘Did it hurt when I got the shot?’ ‘Have I felt unwell?’ anything like that. So, that’s mostly what the questions are. Sometimes I get the, ‘Oh my God. Why would you do that?’ type of situation.”

Some concerns reflected what participants viewed as politically motivated mistrust of COVID-19 mitigation measures. For example, P13 said that “far right” friends from his youth “have made comments like, ‘Well, let me know when you grow your 11th tail.’” More commonly, participants described interlocutors as not inherently mistrustful of vaccines or COVID-19 mitigation, but concerned for participants’ health in the specific case of a COVID-19 trial vaccine. For example, P9 explained that her loved ones were “not anti-vaxxers” but worried since “this [vaccine] is a brand new one, and there’s still so much unknown.” In some cases, relatives feared that side effects would compromise participants’ caregiving ability. For instance, P3 said that “my parents both tried to talk me out of it” because they thought “let somebody else be the first, you’ve got a child to raise, you’ve got us to take care of.”

Participants used a few key responses to allay these concerns. They sought to contextualize the trial COVID-19 vaccine among pre-existing, commonly accepted vaccines. For example, P13 responded to the fears about his “11th tail” by saying “And then you have to have the conversation like, or did you have your measles, mumps, rubella or did you get your polio vaccine? I mean, why don’t you believe in this one, but you believe in the other ones.” Participants often compared the trial COVID-19 vaccine to the flu shot. P2 reported saying that “It was just like the flu shot or any other vaccine,” while P12 explained to others that the side effects he experienced were “similar to what people might get from a flu shot when they have an autoimmune response.”

Others sought to minimize and contextualize the potential risks of vaccine side effects by comparing them to the harms of COVID-19. For instance, P12 said, “I have gotten a question or two about, ‘How bad are the side effects?’ and my response is always, ‘Better than a ventilator.’” Some reframed side effects as a desirable indication of immune response rather than as sickness. P7 noted, “people think they’re getting sick from [vaccination]. They’re not. Their bodies are just reacting to that antigen.” Others allayed concerns by explaining the science behind the vaccine. P25 described such a conversation with concerned friends, explaining, “I guess they don’t really know how vaccines work, and so they were concerned that I could get sick from the vaccine, but I explained to them you can’t.” P6 used humor to turn co-workers’ concerns into “a running joke.” He recalled that a colleague said, “I don’t know how you can do that. What if it turns your brain into a zombie?” When she texted to ask how his first vaccination had gone, he replied, “I feel pretty good but I’m starting to get a craving for brains.” He explained that such joking had diffused tensions about the topic in his workplace.

The second common concern participants reported hearing regarded COVID-19 vaccine development, specifically that an overly fast process and/or one compromised by political interference could lead to a harmful vaccine. Participants responded to fears about fast development by reassuring others that the vaccine had undergone standard testing within a trustworthy institutional context. For example, when P26’s sibling voiced the concern that “I don’t trust the Trump administration not to put a vaccine into phase three trials too soon,” she sought to de-politicize the development process by stating, “there’s lots of bureaucrats at the FDA and stuff who’ve been there for ages, and I don’t think all those people would approve it just because Trump wanted them to.”

Participants also sought to frame rapid vaccine development as the outcome of long-term scientific advances and the unique pandemic context rather than rushed science or poor oversight. For instance, P27 reported that many people she told about her trial participation said they wanted to wait to see if the vaccine turned out to be safe. She said, “I think people may think the race to a vaccine may be politically motivated and not as safe because they’re going very quickly through the processes. And one of the things that I tell people is it’s rare to have this many subjects this quickly when you’re racing for a vaccine. So I think it’s a different time when you’re dealing with a pandemic that I think those steps can be accomplished quicker, because you have more people and more motivation.” P15 used her trial experience as evidence to support a similar claim to concerned others. She noted “I would say that everyone’s worried potentially the trial may be going too fast and that they, the vaccine manufacturers, whomever, will put an unsafe product on the market. But I try to educate people that that’s not what it is. …. I mean, I’m not feeling like I’m getting rushed. They’re not shortening my windows of going back for appointments. They’re not changing any part of the setup of how the trial is to proceed.”

## 4. Discussion

### 4.1 Main findings

Most vaccine trial participants viewed helping to end the pandemic and restore normalcy as a societal duty. Trial participation attracted many because of their health or demographic status, support for science, desire to protect themselves and vulnerable others, and wish to counter both vaccine hesitancy and the politicization of COVID-19 vaccines by modeling vaccine acceptance. Few, mostly young, participants identified compensation as the main reason for participating.

In keeping with the desire to promote COVID-19 vaccination, interviewees often told others about trial participation. While a few told only people they expected to be supportive, many shared widely and reported hearing both support and concern. The most common concern regarded adverse effects. Participants responded by comparing their experiences of vaccination and side effects with that of flu vaccination, contrasting side effects with worse possible harms of COVID-19, communicating vaccine science, and joking. Participants frequently heard concerns about the rapid development of the trial vaccine, in terms of the perceived safety risks of rushed testing and/or politically motivated insufficiencies of oversight. Interviewees responded by framing their trial experiences as evidence for the safety and rigor of the vaccine development process and explaining how unprecedented pandemic conditions and scientific advances enabled fast yet safe vaccine development.

These findings likely reflect both the general experience of the pandemic and specific elements of local context. Desires to help, to protect oneself and others, and to end the pandemic might transcend borders. Participants’ emphasis on “normalcy” reflects the intense abnormality of life amid uncontrolled COVID-19 community spread. Relatedly, critiques of politicization reflect the intense politicization of COVID-19 mitigation measures in the US, exemplified by Iowa’s lack of mask mandates. The local ethos of “Iowa nice” - fostering harmony by downplaying difference and avoiding disagreement - likely also shaped interviewees’ critiques of divisive politicization [29].

### 4.2 Implications for COVID-19 vaccine promotion

These findings from trial participants’ lived experience are consonant with the concerns anticipated by US survey respondents to approved COVID-19 vaccines [4]. Findings underscore the need for vaccination promotion campaigns in the US and similar contexts to address key concerns voiced by interviewees’ interlocutors: fear of vaccine side effects and fear that the rapid vaccine development process compromised safety for the sake of speed or due to political interference.

Interviewees’ motivations for trial participation and responses to others’ fears suggest possible responses to these concerns: framing vaccination as a way to protect oneself and others, help others or do one’s duty, and restore normalcy/end the pandemic; using early vaccine recipients’ experiences to demonstrate safety and model vaccine receipt—especially for members of the demographic groups to which those vaccine recipients belong; normalizing COVID-19 vaccines and their possible side effects by comparing them to those of more widely accepted vaccines and minimizing side effects by comparing them to the known possible harms of COVID-19; and explaining how the unique pandemic context and current vaccine science enabled the rapid yet safe development of COVID-19 vaccines as well as their thorough testing. Findings also illustrate the need to tailor promotional materials to local context [15].

### 4.3 Strengths and Limitations

A key strength of this study is its use of the vaccine trial context to generate early findings about the lived experiences of potential COVID-19 vaccine recipients before vaccine approval. This is a useful context for identifying experienced rather than hypothetical attitudes toward a new vaccine, and especially relevant in this case since the public appears to view even approved COVID-19 vaccines as somewhat experimental given their novelty.

This context also poses limitations. That interviewees were receiving an experimental vaccine likely compounded others’ concerns, and the possibility of receiving placebo or ultimately ineffective vaccine made participants’ experiences differ from those of approved vaccine recipients. Additionally, this was a small exploratory study representing a subgroup of the subgroup of people willing to participate in a vaccine trial; findings might not represent the experiences of all trial participants. While ongoing research with diverse populations throughout the vaccine roll-out will be necessary for creating effective promotion materials, the present findings offer an early look at key themes likely to be relevant for similar populations.

## 5. Conclusion

Interviews with US COVID-19 vaccine trial subjects enabled early identification of themes relevant for vaccine promotion. These include vaccination as helping restore normalcy; protecting oneself, others, and groups to which one belongs; normalizing and contextualizing possible side effects; de-politicizing the vaccines and their development; and explaining how the rapid development process was nevertheless safe. In sum, trial subjects’ stated motivations for participating and their reports of others’ concerns offer specific approaches for tailoring promotional messages.

## Data Availability

The data referenced here is not publicly available.

## Declaration of Competing Interests

The authors declare that they have no known competing financial interests or personal relationships that could have appeared to influence the work reported in this paper.

## Acknowledgements

We thank the participants who graciously volunteered to be interviewed. We are also grateful to trial PI Dr. Patricia Winkour for providing access, and to Michelle Rodenburg and the trial staff for their assistance.

## Authors’ Contributions

Emily Wentzell is principal investigator, conceptualized the research project, obtained research site and IRB approval, and secured funding. Emily Wentzell and Ana-Monica Racila performed literature review, interviews, coding, data analysis, and write-up, and approved the final manuscript.

## Notes

### Competing Interest Statement

The authors have declared no competing interest.

### Author Declarations

University of Iowa IRB, ID# 202008019

